# Deep Learning Approach to Parse Eligibility Criteria in Dietary Supplements Clinical Trials Following OMOP Common Data Model

**DOI:** 10.1101/2020.09.16.20196022

**Authors:** Anusha Bompelli, Jianfu Li, Yiqi Xu, Nan Wang, Yanshan Wang, Terrence Adam, Zhe He, Rui Zhang

## Abstract

Dietary supplements (DSs) have been widely used in the U.S. and evaluated in clinical trials as potential interventions for various diseases. However, many clinical trials face challenges in recruiting enough eligible patients in a timely fashion, causing delays or even early termination. Using electronic health records to find eligible patients who meet clinical trial eligibility criteria has been shown as a promising way to assess recruitment feasibility and accelerate the recruitment process. In this study, we analyzed the eligibility criteria of 100 randomly selected DS clinical trials and identified both computable and non-computable criteria. We mapped annotated entities to OMOP Common Data Model (CDM) with novel entities (e.g., DS). We also evaluated a deep learning model (Bi-LSTM-CRF) for extracting these entities on CLAMP platform, with an average F1 measure of 0.601. This study shows the feasibility of automatic parsing of the eligibility criteria following OMOP CDM for future cohort identification.

## Introduction

The use of dietary supplements (DS) has increased over the past two decades with the growing interests in improving overall health.^1,2^ DS use is more prevalent in the older population than younger adults, among women than among men, and the prevalence of use increases with age in both men and women.^2^ The DS users opt to use DS by personal choice and are usually non-smokers, have a lower body mass index (BMI) and exercise regularly.^3,4,5^ The most commonly used DS are multivitamins, omega-3 or fish oil, vitamins B6, B12, C, A and E, iron, selenium, chromium, zinc, magnesium-containing compounds, calcium and calcium-containing antacids.^6,7,8^

Clinical trials are one of the most valuable resources for healthcare practitioners to practice evidence-based medicine as clinical trials are usually accepted as the most unbiased measures of efficacy and safety for new interventions.^9,10^ Patient recruitment is an essential part of the clinical trial with eligibility criteria (study specific patient characteristics) determine whether a patient should be included or excluded from the study.^11^ However, patient recruitment is a challenging and pressing issue for researchers as it has several barriers, including the lack of patient awareness of clinical trials and access to trials, age limitations, complex study designs, fewer eligible patients than expected due to restrictive eligibility criteria and several other reasons.^12,13,14^ An analysis of registered trials showed that approximately 85% of trials were not able to complete required recruitment in the pre-defined time and around 20% of the trials were closed or terminated early due to inadequate patient recruitment,^15^ limiting the statistical power of the evidence related to the new interventions.^16^ Moreover, more than 70% of clinical trial generalizability assessment studies reported low generalizability of completed trials, partly due to low enrollment.^17^

The rapid growth of the electronic health records (EHR) provides an unprecedented opportunity to harness its data to full potential for secondary use.^18^ Moreover, the last few years have also witnessed an increasing number of clinical research networks focused on building large collections of data from EHRs and claims to provide cohort discovery services. Two notable examples are the National Patient-Centered Clinical Research Network (PCORnet), funded by Patient-Centered Outcomes Research Institute (PCORI), and the CTSA Accrual to Clinical Trials (CTSA ACT) initiative.^19,20^ In addition, a number of national efforts are building tools, algorithms, and data models to identify the eligible patients and to reduce the recruitment delays due to the aforementioned challenges. For example, (1) i2b2 has a widely used cohort discovery tool^21^; (2) the Electronic Medical Records and Genomics (eMERGE) Network is building computable phenotypes for cohort discovery^22^; (3) the stakeholders of the Observational Health Data Sciences and Informatics (OHDSI) consortium are developing open source analytical tools based on the OMOP (Observational Medical Outcome Partnership) Common Data Model (CDM).^23^ The majority of the approaches develop computable representations of the clinical trial eligibility criteria and apply it to EHR data to find eligible patient cohorts. However, as the eligibility criteria is majorly in primarily in free-text format, it is essential to understand the schema of the criteria, the elements (entities and attributes) and potential to parse the data to extract the elements to provide decision support for clinical trial cohort identification.^24^

The NER approaches such as EXACT^25^, EliXR^26^, EliIE^27^, ULTRA^28^, etc., which were developed to represent eligibility criteria in a structured format and were confined to certain drugs and medical conditions. Recently, Si et al. developed a natural language processing (NLP) system to extract medical terms in eligibility criteria of Alzheimer’s disease clinical trials and represent them using the OMOP CDM.^29^ Criteria2Query was developed to systematically transform eligibility criteria text into SQL queries over OMOP CDM databases.^30^ To the best of our knowledge, understanding the eligibility criteria of DS trials and developing computable representations have not been investigated. From our previous published study, we identified that the dietary supplement (DS) trials eligibility criteria are different from drug clinical trials in the aspects like trial objectives and criteria related to trial objectives, demographics (such as age, gender, race), and disease or lab parameters. For certain diseases, drug clinical trials are more therapeutic oriented whereas dietary supplements are either preventive and therapeutic. ^31^ Thus, making the DS clinical trial eligibility criteria unique. So, the objective of this study was to (1) understand data elements associated with DS trials’ eligibility criteria and assess if they can be mapped to OHDSI CDM; (2) develop and evaluate NLP methods, especially deep learning-based models, for extracting eligibility criteria data elements. In this project, we first manually annotate free-text eligibility criteria from a sample of 100 DS clinical trials following OMOP CDM v6.0 and then train and compare both conventional machine-learning-based versus deep-learning-based models on the CLAMP platform to automatically extract different components of eligibility criteria.

## Background

### Observational Medical Outcomes Partnership Common Data Model (OMOP CDM)

The OMOP CDM harmonizes the disparate observational databases with minimal loss of information and enables the interoperability among the databases.^23^ It aims to facilitate research, support the conduct of EHR and manage the claims data. An integral part of CDM is OMOP standardized vocabularies which enable the exchange of patient data among different systems and allow mapping for use in research (https://ohdsi.github.io/TheBookOfOhdsi/). In this study, the definition of entities and attributes such as observation, procedure, device, condition, drug and measurement, are determined following the OMOP CDM standardized vocabularies and clinical data tables.

### Named Entity Recognition (NER)

Clinical NER is a critical task for information extraction (IE) from text and to identify semantics (for example, entities, attributes, relations and events) to support the clinical and translational research. The widely-used clinical NER approaches include MedLEE^32^, MetaMap^33^, KnowledgeMap^34^ and cTAKES^35^. These systems were designed for clinical notes in EHRs or text in the biomedical literature. Studies that investigate the adaptability of these systems to parse clinical trial eligibility criteria are limited. EliXR^26^, EliIE^27^ developed by Weng et al. and EXACT^25^ developed by Yu et al. were designed specifically for parsing eligibility criteria of drug clinical trials. These tools are often limited to certain types of entities and attributes. Early NER methods used dictionary-based, rule-based and supervised machine learning. Later, hybrid methods became popular as combinations of different methods improved the overall performance. Most recently, state-of-the-art deep learning methods have been widely used in textual data representation. One of the most effective NER models being used is Bi-directional long short-term memory (LSTM) with a Conditional Random Field (CRF) on the top layer (Bi-LSTM-CRF).^36^

### Clinical Language Annotation, Modeling and Processing (CLAMP)

CLAMP is a clinical language annotation, modeling and processing software designed by Xu et al.^37^ It is a user-friendly tool and follows the Unstructured Information Management Architecture (UIMA) architecture. The key building blocks of CLAMP are NLP pipelines, machine learning and hybrid approaches, corpus management and annotation tool. CLAMP has multiple components such as sentence boundary detection, tokenizer, part-of-speech tagger, section header identification, abbreviation recognition and disambiguation, named entity recognizer, assertion and negation, UMLS encoder and rule engine. CLAMP’s named entity recognizer contains different types of NER approaches - machine learning-based, dictionary-based, regular expression-based, and deep learning-based. The software has diverse applications in different clinical domains.^38^

## Methods

### Overview of the study

In this study as shown in **Figure 1**, we followed five steps: (1) obtaining the clinical trial eligibility criteria of DS clinical trials from ClinicalTrials.gov; (2) analyzing the eligibility criteria and mapping to OMOP CDM v6.0; (3) developing the gold standard annotation; (4) developing named-entity recognition algorithms (i.e., CRF and Bi-

**Figure 1.**
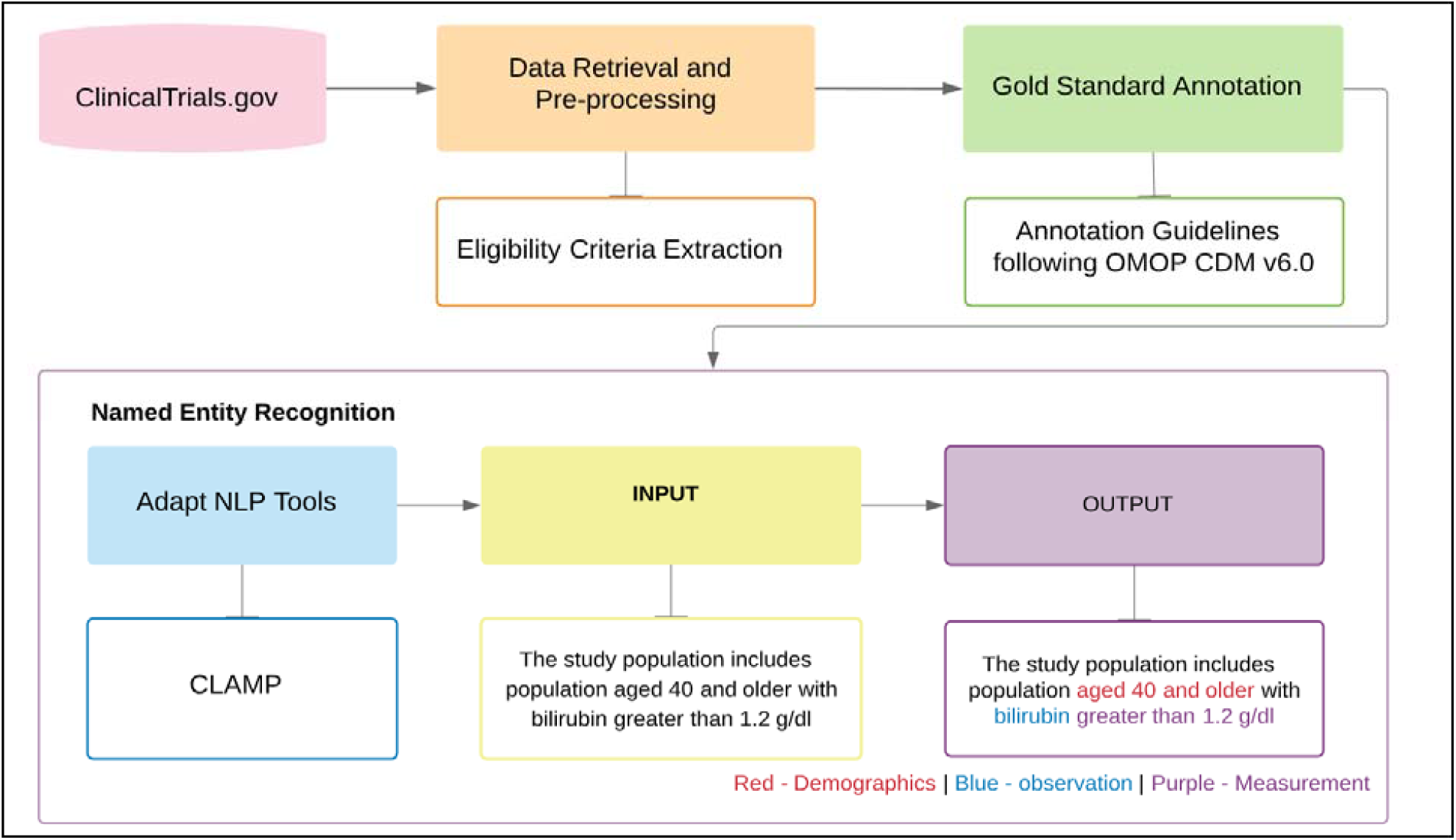
Overview of the Method for Extracting Clinical Trial Eligibility Criteria. LSTM-CRF) using CLAMP; and (5) evaluating the NER models using gold standard annotations.

### Data source and collection

ClinicalTrials.gov is an online repository developed by the U.S. National Library of Medicine (NLM) and the National Institutes of Health (NIH). The repository contains 332,005 research studies which are privately or publicly funded.^39^ We obtained 859 clinical trials from ClinicalTrials.gov by applying the search criteria: (1) using DSs as an intervention, (2) NIH funded trials, and (3) restricting the location to the United States. The extracted trials belong to 22 disease categories. We limited the study to *Behaviors & Mental Disorders and Nervous System Diseases* consisting of 149 trials and *Nutritional and Metabolic Disorders* comprising 199 trials because these two categories of disease categories are the most prevalent domains. Clinical trial data can be obtained in various forms – XML, PDF, plain text, etc. We chose the XML format as it retains the structure of the original document. The document contains both the structured and unstructured data clearly marked with the respective section tags as shown in **Figure 2**. The documents were parsed to extract the eligibility criteria.

**Figure 2.**
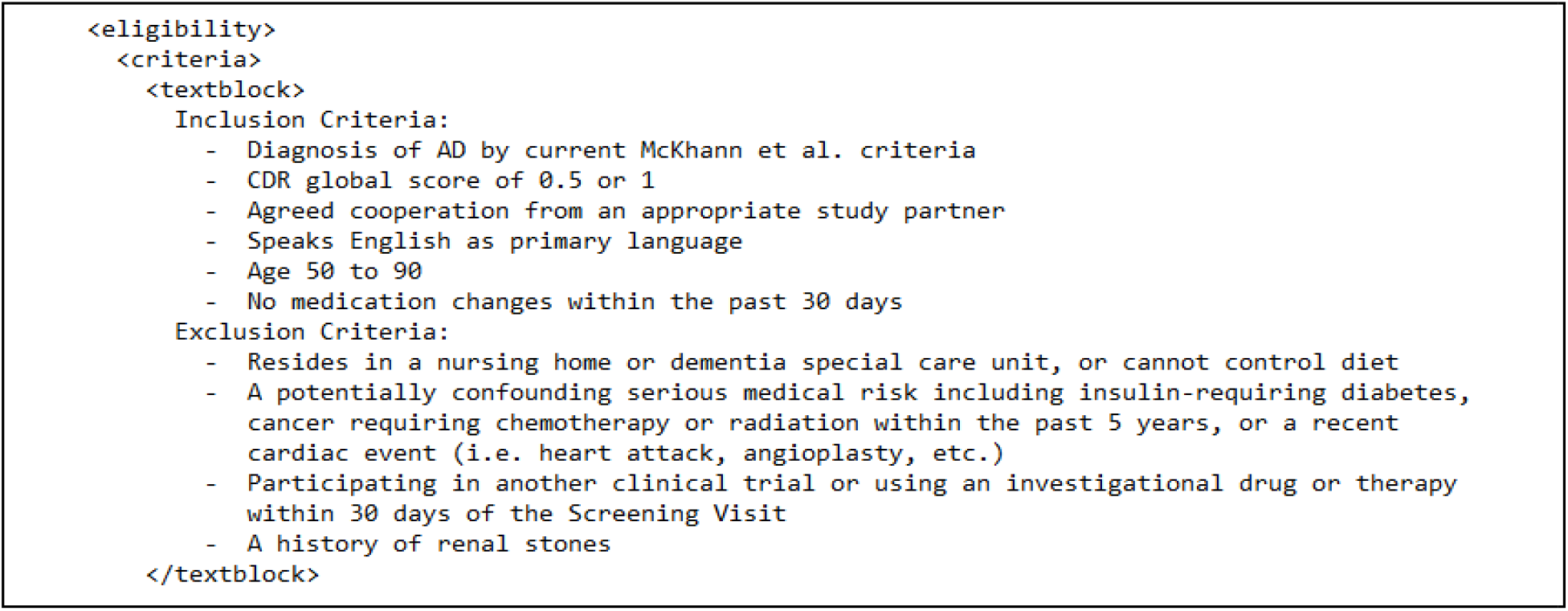
The eligibility criteria section of an example clinical trial (NCT03860792)

### Analyzing the eligibility criteria and mapping to OMOP CDM

Eligibility criteria, including both the inclusion and exclusion criteria, are the list of requirements that an individual must satisfy to be enrolled in the clinical trials. Eligibility criteria can be either short or lengthy, largely free text descriptions spanning several sentences. Each trial comprises an average of 10 criteria (including inclusion an exclusion). Eligibility criteria contain the information about the individual demographics, observation and findings, condition, lifestyle and treatment, as shown in **Figure 3**. The temporal measurement, which is essential but not in the schema, is an element associated with observation, diagnosis, prognosis and treatment. As OMOP CDM standardizes data using a common information model and multiple standard terminologies bridging the interoperability among disparate observational databases, we compared the elements in the schema with OMOP CDM v6.0, domains and mapped the elements to OMOP CDM data tables. We observed that entities like condition, observation/findings and lifestyle, procedures, demographics from the schema can be mapped to OMOP CDM data tables such as condition, observation, procedure and person, respectively. Whereas the sub-entities in the treatment such as drug and device can be mapped to drug and device in OMOP respectively. We found that information about dietary supplements is missing from the OMOP CDM data tables and this element makes this study unique.

**Figure 3.**
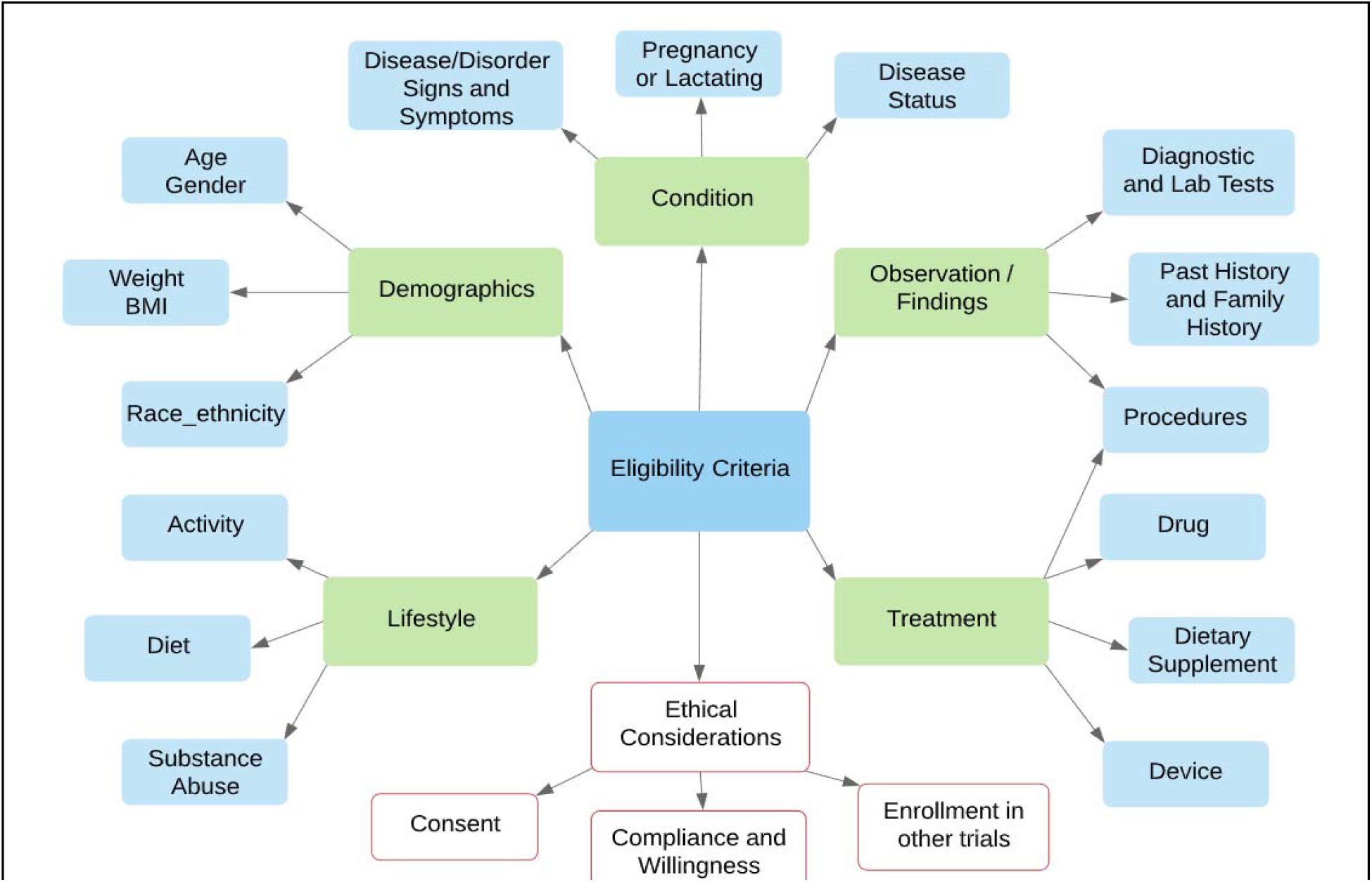
Schema of the elements in clinical trial eligibility criteria. In this figure, different colors are used for the main categories (green) as well as the components (light blue) in eligibility criteria. The category ‘ethical considerations’ and the components associated with it are marked in red as they cannot be found in EHR. The schema laid the foundation to map entities to OMOP CDM.

### Manual annotation

We developed the first iteration of the annotation guidelines based on the OMOP CDM v6.0. We added dietary supplements as an entity as it appears in these DS clinical trials. Three annotators (AB, YX and NW) independently annotated 5 randomly selected trials using CLAMP by understanding the first iteration of the guidelines. The team compared the annotation results, discussed the difference of opinions and revised the annotation guidelines. While annotating, we observed that certain criteria have information about lifestyle choices and annotated the lifestyle choices as observation following OMOP CDM. Next, the team independently annotated 5 trials from each category. The team then discussed and annotated another set until a reasonable interrater agreement is reached and until no discrepancy among annotators. Inter-annotator agreement was computed over 10 trials, revealing a kappa of 0.94. After finalizing the annotation guidelines^a^, totally 100 trials, 50 trials for each of the two categories, were randomly selected for final corpus annotation which comprised approximately 1843 Sentences. The entities and attributes described in the guidelines are given below in **Table 1**.

**Table 1.** Eligible criteria entities and attributes with selected examples.

|  | Semantic Class | Example Criteria (entities and attributes are underlined and marked in blue) |
| --- | --- | --- |
| Entity | Demographics | <u>Women</u> must be > <u>18 to 45 years of age</u>; BMI = 27 kg/m2; |
|  | Observation | <u>Bilirubin</u> greater than 1.2 g/dl; <u>MMSE</u> below 24, dementia or unstable clinical depres |
|  | Procedure | History of <u>bilateral hip replacement</u> |
|  | Condition | Uncontrolled <u>hypertension</u> (BP over 180mm HG) |
|  | Drug | Taking <u>metformin</u>, <u>propranolol</u> and other <u>medications</u> |
|  | Dietary supplement (ds) | Use of <u>St. John's Wort</u> or any other <u>dietary supplement</u> |
|  | Device | Claustrophobia, <u>metal implants</u>, <u>pacemaker</u> or other factors affecting feasibility and / or safety of MRI scanning |
| Attribute | Measurement | BUN <u>above 40 mg/dl</u>, Cr <u>above 1.8 mg/dl</u>, CrCl <u>< 60 mg/dl</u> |
|  | Qualifier | Signs and symptoms of <u>increased</u> intracranial pressure; <u>severe</u> hypercalcemia |
|  | Temporal_measurement | Use of systemic corticosteroids <u>within the last year</u> |
|  | Negation | Use of anti-diabetic drugs <u>other than</u> metformin |

### Developing Named Entity Recognition algorithms using CLAMP

We used CLAMP as a platform to develop NER algorithms on 1843 annotated sentences. We chose CRF as the baseline model. We also implemented a deep learning model, Bi-LSTM-CRF model, using TensorFlow framework, which has been demonstrated superior performance in other NER tasks.^36^ LSTM network is better than traditional RNNs to find long range dependencies due to their updated hidden layer. As NER tasks often require contextual information (both past and future input features) from the sentence, we made use of a bidirectional LSTM network. The Bi-LSTM networks were trained using backpropagation through time technique and both forward and backward hidden states were concatenated to obtain contextual representations for the input sentence. To make full use of contextual tagged information, we then combined the Bi-LSTM networks with a CRF network to get a Bi-LSTM-CRF network. Finally, a 5-fold cross validation (80 trials for train and 20 trials for test) was applied to compare the performances of two models. The NER performances on each entity and attribute were reported using precision, recall and F1-measure.

## Results

### Distribution of dietary supplements in clinical trials

In this study, we observed that a wide range of DS have been studied as the interventions in clinical trials. Figure 4 lists the distribution of trials on each DS for two categories of diseases. As shown in Figure 4, the most studied dietary supplements in trials on Behaviors & Mental Disorders and Nervous System Diseases were fish oil, omega-3 fatty acids, vitamin D, vitamin E, DHA, EPA, soy, lipoic acid, selenium, folic acid and the those only studied once were black cohosh, boswellia serrata, chamomile extract, etc.

**Figure 4.**
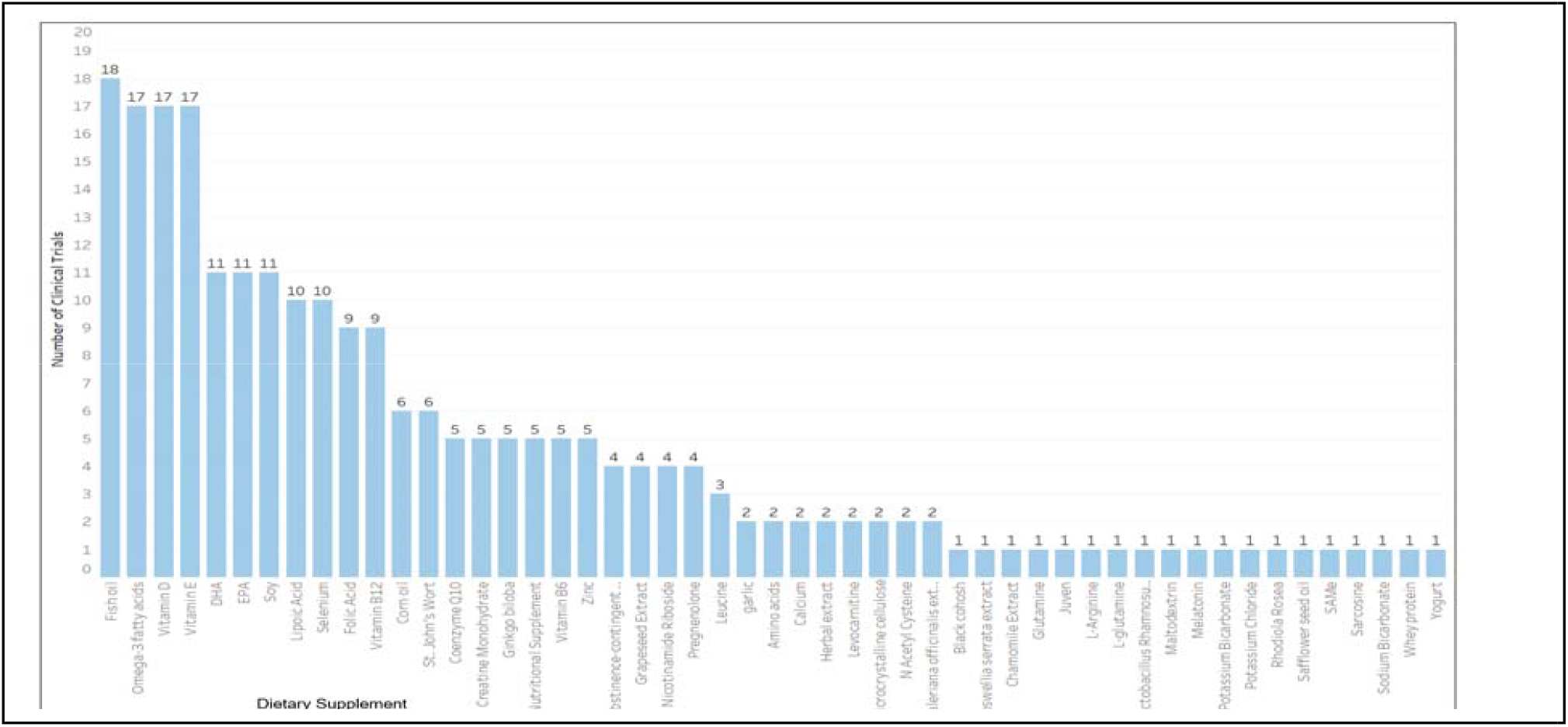
Distribution of DS as intervention in clinical trials on Behaviors & Mental Disorders and Nervous System Diseases Whereas in those trials on Nutritional and Metabolic Diseases (**Figure 5**), vitamin D is predominated with 130 studies, followed by calcium, fish oil, omega-3 fatty acids, chromium picolinate were widely studied.

**Figure 5.**
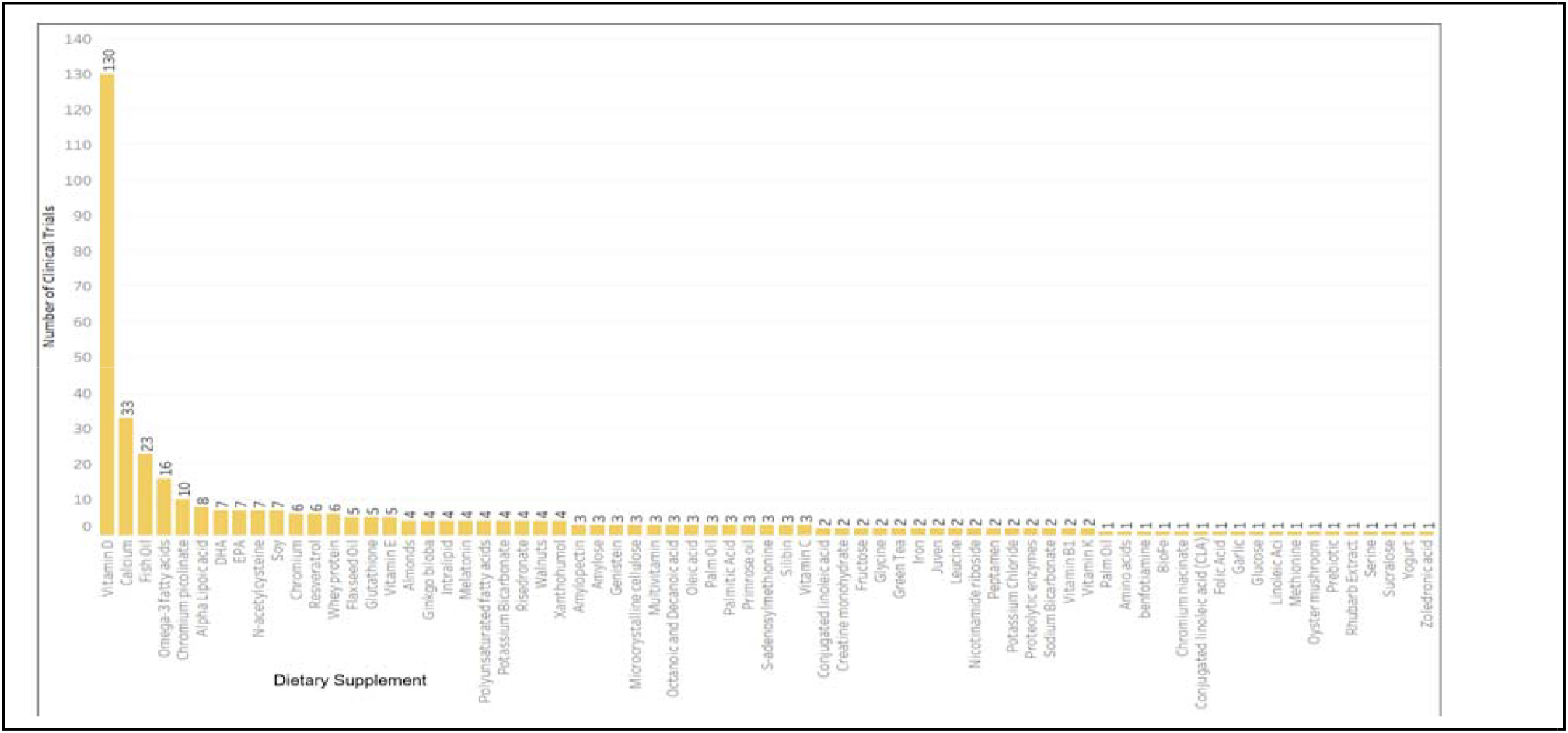
Distribution of DS as intervention in clinical trials on Nutritional and Metabolic Diseases

### Statistics of Eligibility Criteria

Among the 1843 eligibility criteria sentences, 229 criteria were not annotated as the criteria were not computable (corresponding data not in EHR). Descriptive statistics of the unannotated and annotated eligibility criteria corpus are provided in **Table 2** and **Table 3**, respectively. Among the unannotated 229 sentences, 23.14% of the sentences belong to the criteria referring to unwillingness or willingness of the subject, whereas 2.18% belong to partners or caregivers. Out of annotated 1614 sentences, condition entity is the largest (1401 terms), followed by drug (688 terms) and observation (671 terms) while device is the smallest (47 terms). Among the attributes, the qualifier is the largest (643 terms) followed by temporal measurement (445 terms). The average of terms of each semantic class that can be found in a trial range approximately from 2 to 5.

**Table 2.** Descriptive statistics of the unannotated eligibility criteria and examples.

| Category | No. (%) | Example Criteria |
| --- | --- | --- |
| Unwillingness / Willingness | 53 (23.14) | unwillingness for subject of childbearing potential to use contraception during the first year of the study |
|  |  | She must be willing to practice an acceptable method of birth control |
| Others | 39 (17.03) | The patient does not have an outside care provider for treatment of depression |
|  |  | Access to a smart phone or internet and telephone |
| Ability / Inability | 31 (13.54) | Inability to swallow oral capsules |
|  |  | Ability to use a bolus calculator function with the current insulin pump with pre-defined parameters for glucose goal, carbohydrate ratio, and insulin sensitivity factor |
| Informed consent | 28 (12.23) | Signed protocol specific informed consent prior to registration |
|  |  | Subjects who refuse to sign the protocol consent document |
| Participation in other trials | 25 (10.92) | Enrollment in any concurrent research protocols that would interfere with participant safety or research data integrity |
|  |  | Previous participation in Phase 1 pharmacogenomic study |
| Investigator's opinion | 21 (9.17) | The patient is, in the opinion of the investigator, mentally or legally incapacitated |
|  |  | Any other reasons that, in the opinion of the Investigator, the candidate is determined to be unsuitable for entry into the study |
| Location | 14 (6.11) | Lives far away from study site |
|  |  | Live and work within 1 hour of the study site |
| Language | 13 (5.67) | Fluency in English or Spanish |
|  |  | Non-English speaking |
| Partners/care takers | 5 (2.18) | Both parents/partners are required to participate in this study, not just one or the other |
|  |  | Living at home with a parent or guardian |

**Table 3.**
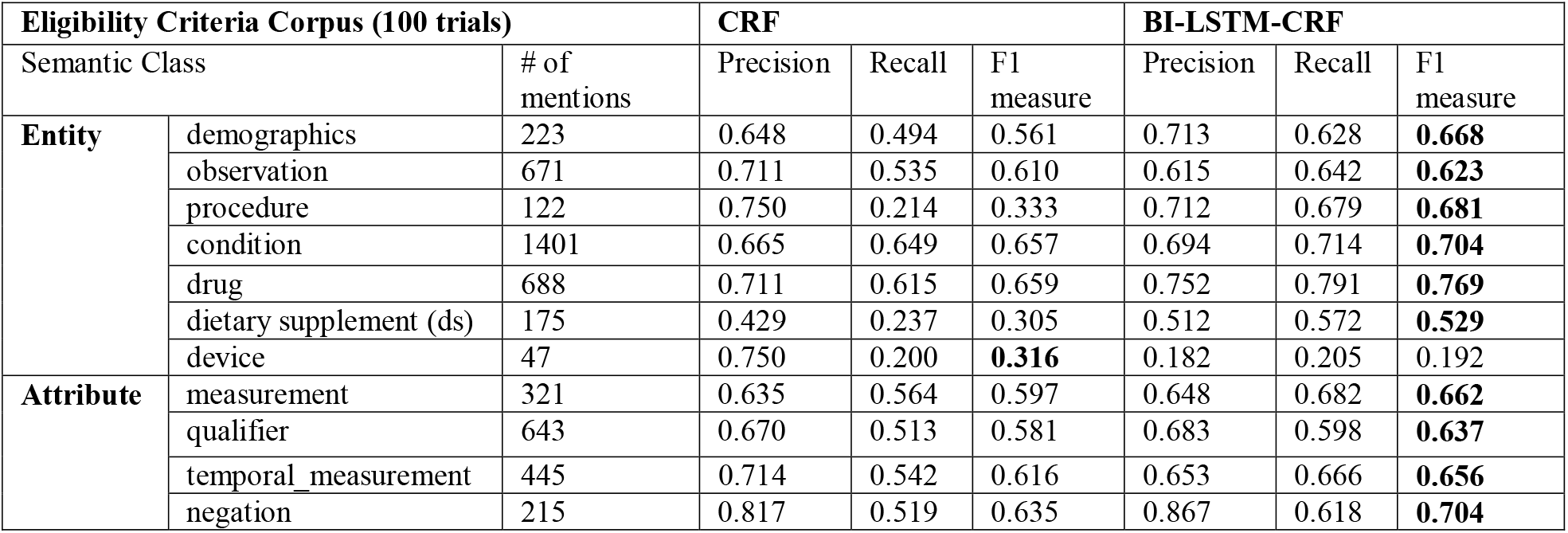
Descriptive statistics of the annotated eligibility criteria and performance of the models for entity and attribute recognition. The best F1 measures for each semantic class are in bold.

### NER Models Evaluation

The detailed performance of the CRF and Bi-LSTM-CRF for the named entity recognition task is given below in **Table 3**.

In almost all entities and attributes except device, Bi-LSTM-CRF outperformed CRF model. The macro-average of F1 measure for the Bi-LSTM-CRF model is 0.601. The semantic class which performed the best in the Bi-LSTM-CRF model is drug with an F1 measure of 0.769 followed by condition with 0.704. The semantic class which performed the least in Bi-LSTM-CRF is device (0.192).

## Discussion

The widespread adoption and use of EHRs together with the NLP tools have led to the ability to identify and recruit patient cohort to conduct clinical trials according to eligibility criteria. However, EHRs may or may not contain all eligibility criteria data elements required for patient cohort identification. One could also incorporate clinical notes in the EHR to find patients that meet criteria that are not captured in the structured field in EHR, e.g., MMSE of dementia patients. The eligibility criteria schema and elements should be better understood and analyzed to differentiate the criteria that are not computable.

In this pilot study, we first analyzed the eligibility criteria schema and elements (entities and attributes) for the DS clinical trials obtained from ClinicalTrials.gov for two disease categories, *Behaviors & Mental Disorders and Nervous System Diseases*, and *Nutritional and Metabolic Diseases*, the top two categories based on the frequency of DS uses. Eligibility criteria comprise multiple elements which are bound by restrictions such as qualifiers (severity of disease), negations and temporal constraints; identifying the individual entities and attributes would make EHR computability easier. As our long-term goal is to accelerate the patient recruitment using EHR data, we followed the commonly used OMOP CDM to annotate the eligibility criteria. Through our analysis on eligibility criteria of DS clinical trials, we found that dietary supplement is currently out of scope of OMOP CDM. As clinical notes will contain information about dietary supplements, we included the entity in our annotation and NER development. While annotating we observed that certain criteria can be easily computable to extract from EHR data while the rest are either hard or impossible to compute. The eligibility criteria whose corresponding data cannot be found in the EHR were not annotated even if certain terms in the criteria qualify for one of the entities or attributes as this information is not computable.

The annotators faced a few challenges while annotating the criteria. For example, the annotators had noted certain problems while annotating the criteria including (1) the criteria had both the qualifier and disease condition (e.g., multiple myeloma); (2) the criteria were difficult to differentiate as they shouldn’t be annotated (e.g., aerobically trained with V02 max greater than 2 SD above age-adjusted mean); (3) ambiguity (e.g., certain abnormal laboratory values); (4) had interconnected entities (e.g., hepatic, renal and gastrointestinal diseases). The annotators had overcome these challenges through multiple rounds of discussions and annotations until no discrepancy among annotators.

To explore the feasibility of the NLP techniques, especially state-of-the-art deep learning models, for parsing these eligibility criteria automatically, we leveraged the annotation and CLAMP platform to compare two models. The precision and recall of the entities and attributes for the CRF model are mostly lower than the Bi-LSTM-CRF model except for entity ‘device’. In the Bi-LSTM-CRF model, within the 5 folds for the entity ‘device’, three folds did not find the entity whereas the other two showed low precision and recall values. By analyzing the original train/dev/test dataset, we observed that the difference in values in different folds is due to a small dataset for device (totally only 47 mentions). CRF requires a list of features, while Bi-LSTM-CRF is a feature engineering free model but require a large dataset. This is one main reason why the current performance is still suboptimal. The most common semantic classes “drug”, “condition” and “negation” in clinical trials reached the F1 measure over 0.70 whereas the F1 measure for other semantic classes is above 0.50 except for the semantic class “device”. The low F1 measures could be due to a small dataset and lower number of mentions in the annotation corpus.

This study has a few limitations. The major limitation is a small dataset which resulted in low performance with respect to certain entities and attributes. The other notable limitation is the differences in individual perception while annotating certain concepts (e.g., condition, qualifier) in the dataset which resulted in some inconsistent annotations. Future work will focus on using a large dataset and improving the annotations consistency which would eventually improve the performance of the NER models. We will also try other deep learning modes, such as BERT, which shows promising performance in 11 NLP common tasks.

## Conclusions

In this study, we investigated the data elements associated with eligibility criteria associated with the clinical trials which use DS as an intervention. We analyzed the criteria and found both computable and non-computable criteria. We manually created eligibility criteria entities followed the OMOP CDM v6.0. We annotated these entities for 100 trials and used the annotated data to develop a Bi-LSTM-CRF model for NER task. This study demonstrates the feasibility of using CDM to represent the DS clinical trial eligibility criteria and using deep learning models for NER task in clinical trials. This study lays the foundation for future matching patients using their EHR data to DS clinical trials.

## Data Availability

The data pertaining to this article is available upon request.

## Acknowledgements

This work is partially supported by the National Center for Complementary and Integrative Health (NCCIH) and the Office of Dietary Supplements (ODS) under grant number R01AT009457 (Zhang); and the University of Minnesota Clinical and Translational Science Institute (CTSI), supported by the National Center for Advancing Translational Sciences under grant number UL1TR002494.This study was also partially supported by the National Institute on Aging (NIA) of NIH under Award Number R21AG061431; and the University of Florida CTSI, which is supported in part by the National Center for Advancing Translational Sciences under award number UL1TR001427. The content is solely the responsibility of the authors and does not necessarily represent the official views of the NIH.

## Footnotes

a https://z.umn.edu/annotation_guidelines

